# The COVID-19 Pandemic and Early Child Cognitive Development: A Comparison of Development in Children Born During the Pandemic and Historical References

**DOI:** 10.1101/2021.08.10.21261846

**Authors:** Sean CL Deoni, Jennifer Beauchemin, Alexandra Volpe, Viren D’Sa, the RESONANCE Consortium

## Abstract

**Objective:** To characterize cognitive function in young children under 3 years of age over the past decade, and test whether children exhibit different cognitive development profiles through the COVID-19 pandemic.

**Study Design:** Neurocognitive data (Mullen Scales of Early Learning, MSEL) were drawn from 700 healthy and neurotypically developing children between 2011 to 2021 without reported positive tests or clinical diagnosis of SARS-CoV-2 infection. We compared MSEL composite measures (general cognition, verbal, and non-verbal development) to test if those measured during 2020 and 2021 differed significantly from historical 2011-2019 values. We also compared MSEL values in a sub-cohort comprising infants 0-16 months of age born during the pandemic vs. infants born prior. In all analyses, we also included measures of socioeconomic status, birth outcome history, and maternal stress.

**Results:** A significant decrease in mean population MSEL measures was observed in 2021 compared to historical references. Infants born during the pandemic exhibited significantly reduced verbal, non-verbal, and overall cognitive performance compared to children born pre-pandemic. Maternal stress was not found to be associated with observed declines but a higher socioeconomic status was found to be protective.

**Conclusions:** Results reveal a striking decline in cognitive performance since the onset of the COVID-19 pandemic with infants born since mid-2020 showing an average decrease of 27-37 points. Further work is merited to understand the underlying causative factors.

## INTRODUCTION

The outbreak of the SARS-CoV-2 (COVID-19) pandemic in the USA in the latter part of 2019 and early months of 2020 has brought widespread change and disruption to our social, economic, and public health environments. While children have largely been spared the severe health and mortality complications of SARS-CoV-2 infection (1, 2), they have not been immune to the impact of the public health and social distancing policies that closed daycare centers, schools, and playgrounds (3, 4) and limited opportunities for play and interaction. While these policies helped to limit COVID-19 spread they also disrupted educational opportunities (5), limited interaction with other children and caregivers (6), and reduced opportunities for physical activity and play (7). More broadly, these policies have also shuttered businesses and led to large-scale employment shifts which may have affected the home, food, and financial security of children through parental or caregiver job loss or furloughs.

In light of these social and economic challenges, there has been concern amongst early child researchers regarding the impact of the pandemic on infant and early child development and mental health. While there are few past examples of non-conflict-related lock-downs with a similar geographic extent and prolonged timeline, past studies of acute and chronic environmental stress on child development provide important context. There is little doubt that chronic maternal stress, anxiety, and depression throughout pregnancy can impact fetal and post-natal development (8, 9). Maternal stress experience may be associated with increased fetal exposure to stress-related hormones (e.g., cortisol), which can result in differential brain structural and functional development and later cognitive impairments (10-13). For example, past analysis has revealed strong associations between maternal prenatal stress and anxiety related to maternal or paternal displacement and job loss and infant health (birth weight and gestation duration), mortality, temperament, and cognitive development (14). Severe acute stress, such as that associated with natural disasters (e.g., the 1998 snowstorm that affected central Canada, and the 2006 Hurricane Katrina in Louisiana) has been shown to yield long-term cognitive and intellectual consequences in children who were exposed *in utero* (15, 16). Of note, *in utero* exposure to maternal stress appears to more adversely affect the male fetus (17, 18), implying potential sexual dimorphism in stress sensitivity.

In addition to the potential impact on the developing fetus, children exposed to environmental stress and adversity may also be similarly affected. Prolonged stress, such as experienced during wartime or refugee replacement (e.g., the Bosnian war from 1992-1995) is associated with worsened long-term physical and mental health (19). Recent cross-sectional and longitudinal studies of children and adolescents during the early stages of the pandemic have revealed increased stress, anxiety, and depression (20) alongside reduced academic growth in math and language arts (4).

While pre and post-natal stress and adversity can clearly impact the developing child, these effects may be moderated by compounding social and structural factors such as socioeconomic status, access to healthcare, and other social determinants of health (21, 22). The COVID-19 pandemic has disproportionately affected Latino, Black, African American, and lower socioeconomic status (SES) individuals and families (23, 24). Of reported COVID-19 cases in pregnant women, the majority are Latino (42.5%) and Black and African American (26.5%) (25, 26). 46% of hospitalized children are Latino and 30% are Black or African American (27) and are more likely to develop severe illness (28). Likewise, Latino and Black/African American adults are three times more likely to be hospitalized with COVID-19, and 1.9-2.3 times more likely to suffer severe complications or death (29). Adults with incomes below $15,000 are at 2x greater risk of serious illness than those who make $50,000 or more (30) and they have been hardest hit by the closure of non-essential services, employee furloughs, and childcare and school disruptions.

Finally, changes in the children’s home environment may also have a developmental impact. For example, while associated with stress, reduced income from job loss or layoff may affect the ability to provide optimal nutrition and access to healthcare (31). Closed daycare centers, public parks, and gathering spaces can limit explorative play (32), child-child interaction, and environmental stimulation (22). And parents who may have taken on additional jobs may have experienced reduced time to read and interact with their children. All of these factors have previously been associated with variations in cognitive and behavioral development and outcomes (33-35).

Taken together, these past findings strongly imply the likely impact of the ongoing COVID-19 pandemic and related environmental changes and stressors on child health and neurodevelopment. However, the specific impact on infant and pre-school cognitive development remains less clear, with few studies focused on this early age range from 0-3 years (36). However, emerging results are disturbing. For example, findings from a prenatal cohort in New York City recruiting during the earliest stages of the pandemic reveal reduced motor and cognitive development in infants at 6-months of age (37), and children 6 and 12-months of age born during the pandemic in Guangzhou, China, similarly show reduced communication skills compared to historical references (38).

From 2011, the RESONANCE longitudinal study of child health and neurodevelopment has been co-ordinated from the Warren Alpert Medical School at Brown University in Providence. This study includes approximately 1600 caregiver-child dyads who have been continuously enrolled between 0 and 5 years of age since 2010 and have been followed through infancy, childhood, and early adolescence. This cohort, therefore, offers a unique opportunity to examine the potential impact of the COVID-19 pandemic on child health trends in RI and to compare with other trends observed throughout the US.

Like many regions in the United States, RI implemented a series of school closures, public access, and travel restrictions, and limited social gatherings early on in the pandemic to limit virus spread. Elementary schools were closed to in-person instruction from March to September 2020 and many continued to operate fully online or with hybrid in-person learning until January 2021. Daycares were also closed in March 2020 but were allowed to reopen with limited capacity from June 2020 until June 2021. Broader state-wide travel restrictions and stay-at-home orders were enforced from March to May 2020, with many businesses operating with reduced on-site workforces and/or work-from-home options until mid-2021. Indoor and outdoor mask policies were also in place throughout 2020 and 2021 following CDC guidance. Despite being one of the smaller US states (population ∼1 million), RI has recorded ∼154,000 cases of COVID-19 illness and almost 3,000 deaths and mirrored national trends with respect to disproportionate infections and deaths in Hispanic, Latino, and African American communities (39, 40), and lower-income families (41). Thus pregnant individuals and families in RI experienced similar pandemic-related stresses as those elsewhere in the US.

Drawing from our RESONANCE dataset, we used linear mixed-effects models to examine trends in infant and early child neurodevelopment over the past decade, from 2011 to 2021. We find that even controlling for factors including age, biological sex, socioeconomic status, and maternal stress, measures of verbal, non-verbal, and overall cognitive functioning are significantly lower in 2021 compared to past years. These results provide additional supporting evidence that aspects associated with the COVID-19 pandemic, even in the absence of direct SARS-CoV-2 infection, have impacted infant and early child neurodevelopment.

## METHODS

All data were collected in accordance with ethical approval and oversight by the Rhode Island Hospital institutional review board. Informed consent was obtained from all parents or legal guardians.

Between January 2011 and December 2021, a total of 1247 cognitive assessments have been performed on 700 healthy and neurotypically developing children from 3 months through 3 years of age. At the time of enrollment, all children met the following criteria for inclusion: Full-term gestation (38 weeks or greater); singleton pregnancy; no abnormalities on prenatal ultrasound; healthy birth weight (>2500g); no exposure to cigarette smoke, alcohol, marijuana, or illicit substances *in utero*; uncomplicated delivery (e.g., no history of birth asphyxia), no history of gestation diabetes in the mother; no major psychiatric illness in the mother including depression requiring medication in the year prior to pregnancy; no neurological trauma in the infant; and no diagnosed neurological disorder in the infant (e.g., epilepsy). Further, none of the children or pregnant mothers with newborns reported a positive antibody, antigen, or PCR-TR result for SARS-CoV-2 infection over the past year, or had a clinical diagnosis.

For this study, we examined the potential impact of pandemic-related environmental changes in the following groups: 1. 700 children between 3months and 3 years of age assessed between January 1, 2011, and November 30, 2021; and 2. In a comparison of 525 infants 0-16months of age born before the pandemic (n=388) and during the pandemic (n=137). A pictorial overview of all included child assessment timings is shown in **Supplemental Figure 1**, with demographic details provided in **Table 1**.

**Table 1.**
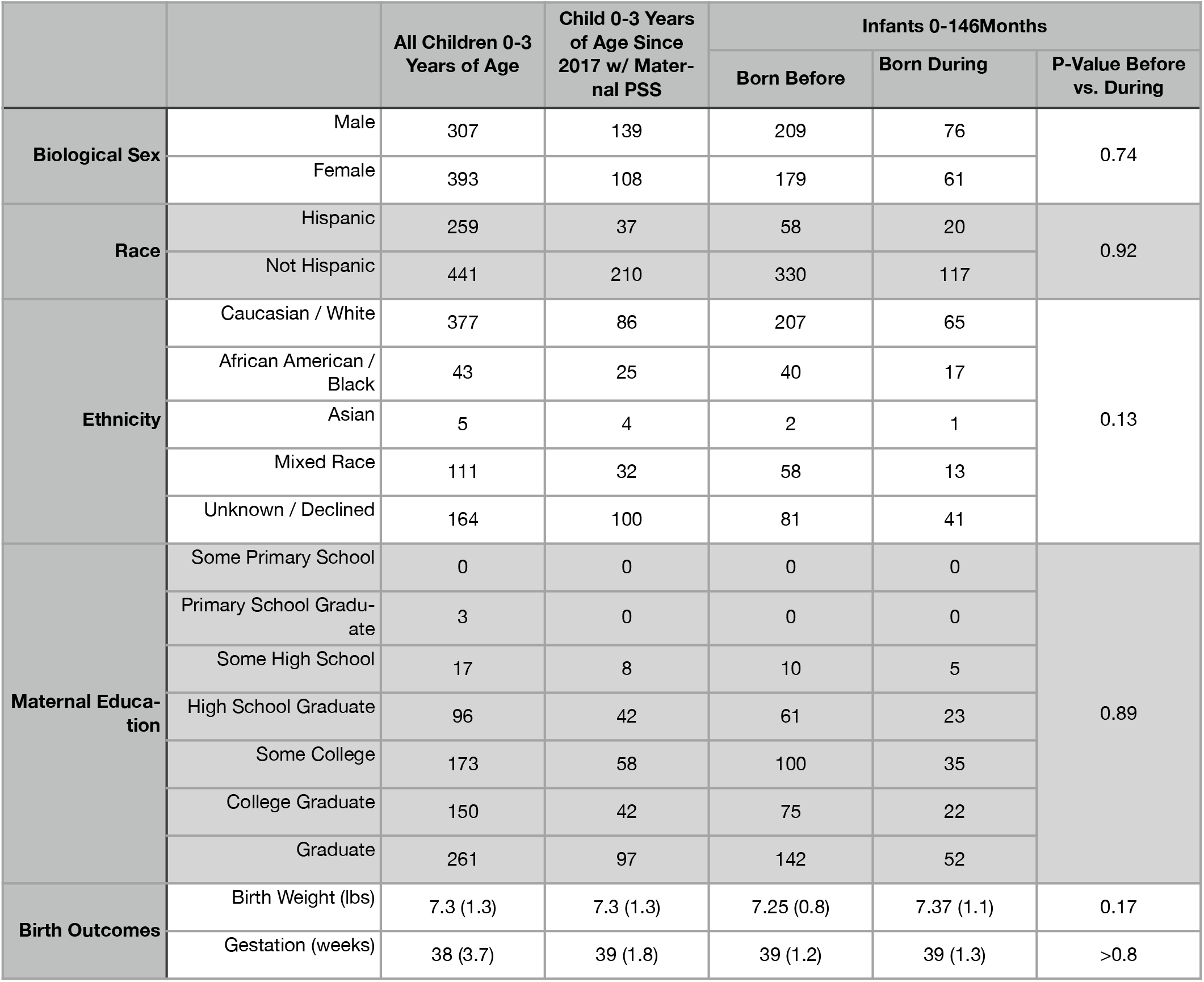
Group demographics for each child cohort, including 1. All 700 children 3-months through 3-years of age tested over the past decade; 2. A subset of 247 children from group (1) with PSS data; and 3. 525 infants 0 to16-months of age born before and during the pandemic We also include the results of chi-square and t-test comparisons between the before and during pandemic infant groups.

### Neurocognitive Assessments

The primary neurocognitive outcome measure assessed in all children was the Mullen Scales of Early Learning, MSEL (42). The MSEL is a population-normed and clinically-administered tool that assesses function across five primary domains: Fine and Gross Motor control, Visual Reception, and Expressive and Receptive Language via direct observation and performance. Raw domain scores can be converted to age-normalized T-scores and combined into the three composite values: the Early Learning Composite, and the Verbal and Non-Verbal Development Quotients (ELC, VDQ, and NVDQ, respectively).

All assessments, both before and during the pandemic were performed in-person in a consistent lab setting and overseen by a consulting neuropsychologist. Following institutional guidelines and directives, in-person assessments during the pandemic were performed with face masks on both staff and participants.

### Demographic, Socioeconomic, and Other Data Collection

Alongside neurocognitive assessments, parent and child health, birth outcome (weight, length, gestation), demographic (race, ethnicity, primary spoken language at home), and socioeconomic data (maternal and paternal education level) were collected via parent report. Highest completed education grade of level was converted to the 7-level Hollingshead scale (43) with 1 = partial elementary school; 2 = elementary school graduate; 3 = partial high school; 4 = high school graduate; 5 = partial college; 6 = college graduate; and 7 = graduate or professional degree.

Since 2017, maternal stress has been measured using the Perceived Stress Scale (PSS) (44), a 10-item self-report that provides a continuous scale of perceived and experienced stress due to life situations.

### Data Analysis

#### Comparison of Pre-Pandemic and Current Mean Cognitive Measures

ELC, VDQ, and NVDQ measures were grouped by testing year from 2011 to 2021. For 2020, we split the sample into two parts corresponding to January 1 to April 1 and April 2 to December 31 - coinciding with the state-wide lock-down and shelter-in-place orders. We then compared measure for each composite score between each pre-post-pandemic year pair (e.g., 2011 vs 2020; 2012 vs. 2020 ……. 2019 vs. 2021) using an analysis of covariance (ANCOVA). Child age and maternal education (as a proxy for socioeconomic status, SES) were included as covariates.

#### Modeling Cognitive Measures over the Past Decade

We further investigated trends in the longitudinal values by constructing a series of general linear mixed-effects beginning with the most simplistic to determine if measures during the pandemic were lower than 2011-2019 values,

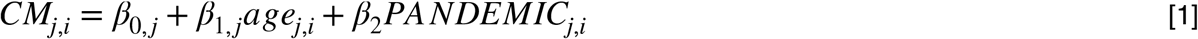

Where CM is the cognitive measure of interest (i.e., ELC, VDQ, or NVDQ) of child *j* at time-point *i. β*_*0,j*_ is the intercept and *β*_*1,j …*.,_ *β*_*n,j*_, are the regression coefficients. *β*_*0,j*_, *β*_*1,j*_ combine a sample fixed effect and a subject-specific random effect to allow individual differences in mean cognitive function and change with age. The PANDEMIC term is a binary factor that is 0 for any testing date prior to March 16, 2020, and 1 for dates thereafter. Equation [1] was fit to the complete cohort dataset using the fitlme function in Matlab (MathWorks, Cambridge, MA v2019b). Missing maternal education, birth weight, and gestation period data were imputed using a random forest algorithm (fitrensemble).

We then used a step-wise approach to systematically include additional factors and interactive terms that may account for observed differences, i.e.,

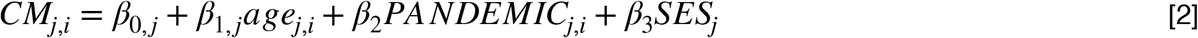

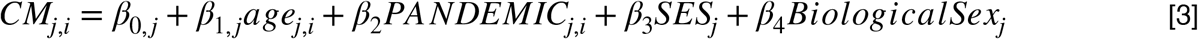

to control for potential differences in development or sensitivity by biological sex and socioeconomic factors. Given past findings associating COVID-19 stay-at-home orders and prematurity or potential low birth weight (45), we further included these birth outcomes as additional predictors,

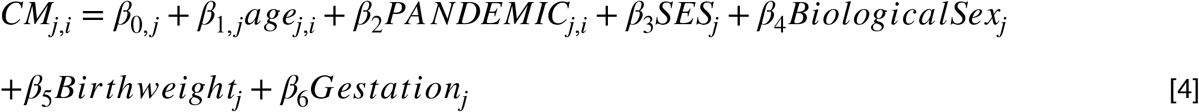

Finally, we also tested the interaction between the PANDEMIC and SES and Biological Sex terms to test whether these factors had additive or buffering effects,

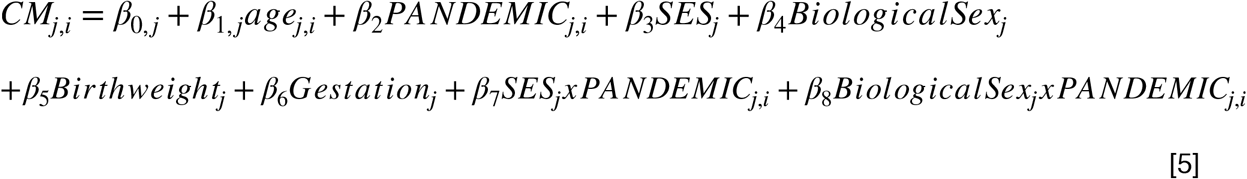

The Bayesian Information Criterion (BIC) (46) was calculated for each model as used as a metric of parsimony. We then examined the significance level of each model parameter, specifically the PANDEMIC term, which denotes a potentially significant difference in pre-and during pandemic scores, in the most parsimonious model.

#### Cognitive Differences in Infants Born During the Pandemic Versus Before

Recognizing that newborns and young infants may be more susceptible to pandemic-related stresses, we sought to specifically investigate the COVID-19 environmental changes on children born since the beginning of the pandemic and lock-down guidelines. Specifically, we performed the same modeling as described above (Eqns. 1-5) on a subset of data that included infants 0-16months of age born before (n=388) and during the pandemic (n=137). (**Table 1, Supplemental Figure 1**).

#### Investigating the Impact of Maternal Stress

In the absence of direct SARS-CoV-2 infection, environmental exposures associated with the COVID-19 pandemic can affect the developing infant and child through multiple pathways. Maternal stress throughout pregnancy is one such mechanism (47, 48). Since 2017, pregnant individuals and mothers have completed the PSS alongside their child’s neurocognitive assessment visit.

We investigated the impact of stress by modifying the most parsimonious model identified from the above analysis by adding PSS as an additional predictor, as well as by replacing the PANDEMIC term with the PSS value in the model, i.e.,

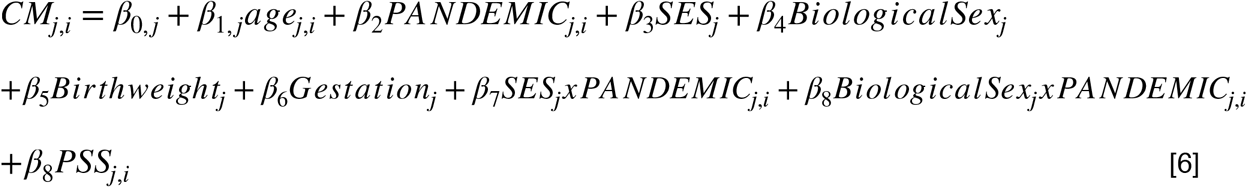

or

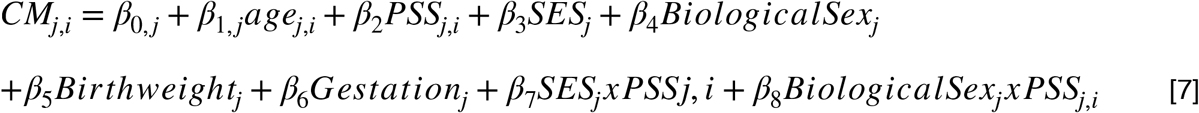

Since the PSS data has only been collected since 2017, data for this analysis was restricted to 247 children, (**Table 1**).

## RESULTS

### Child Demographic and Socioeconomic Profiles

Yearly birthweight, gestational period, maternal education, and maternal PSS scores are visually plotted in **Supplemental Figure 2** and show no significant differences in mean birth weight, gestation, maternal education, or stress profiles of the families and children assessed in 2020/2021 compared with 2011 to 2019.

### Comparing Pre-Pandemic and Current Mean Cognitive Measures

Mean ELC, VDQ, and NVDQ scores for each year from 2011 to 2021 are shown in **Figure. 1** for **(a)** all children between 0-3 years of age; and **(b)** for the subcohort of infants 0-16months of age.

**Figure 1.**
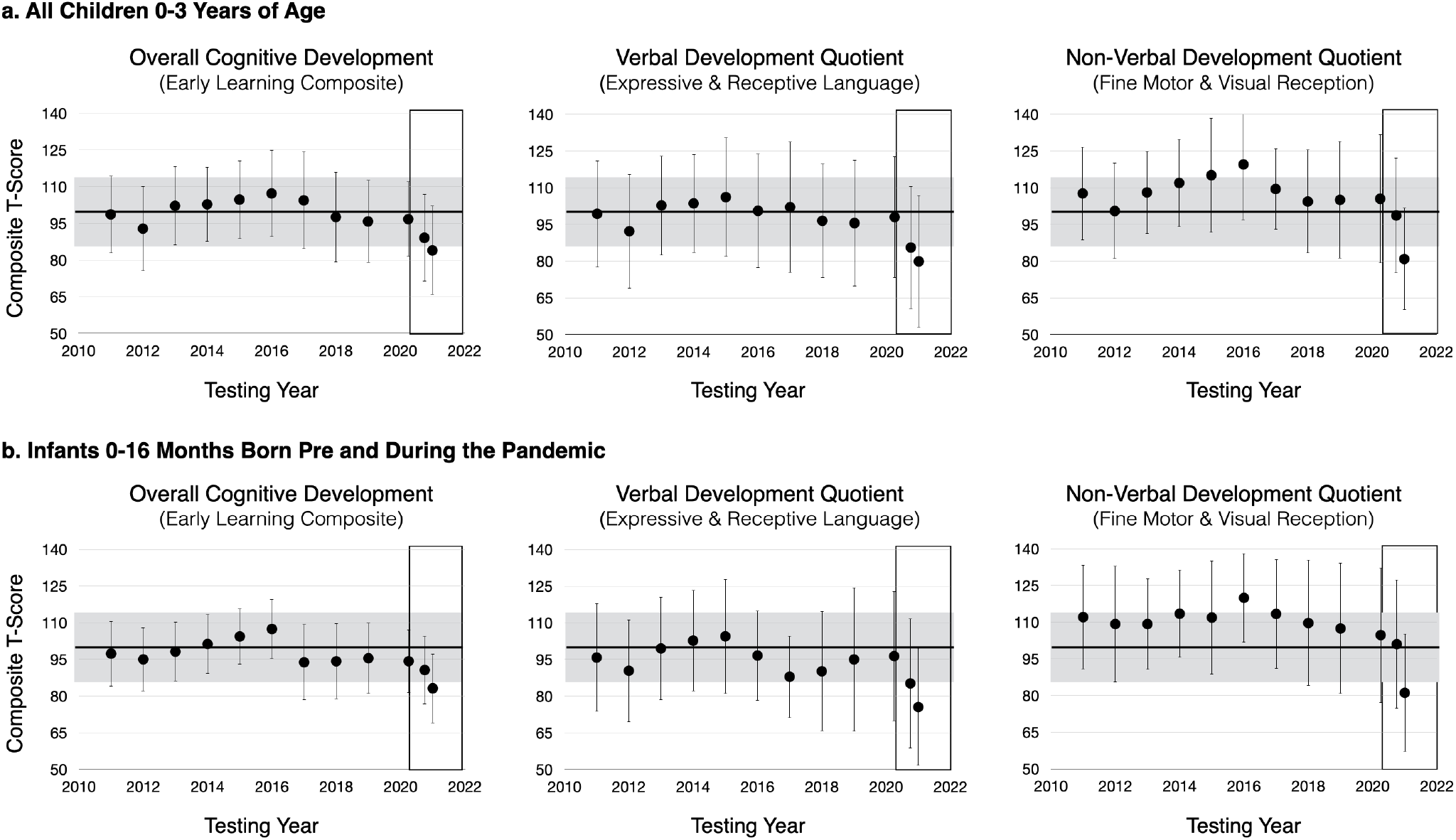
Visual comparison of yearly mean MSEL ELC, VDQ, and NVDQ composite scores for (top) all children 0 to 3 years of age in the cohort; and (bottom) infants 0 to 16 months of age born before or during the pandemic. In each panel, the black line represents the expected mean (100) and the grey region denotes the expected standard deviation (15). For 2020 we provide 2 measures, one for January 1 to April 1, 2020, and the second from April 2 to December 30, 2020. The box denotes measures obtained during the pandemic since April 1, 2020. In both child cohorts, we observe general decreases in cognitive performance in 2021 that, as shown in Table 2, are statistically significant.

In infants and toddlers (**Fig. 1a**) mean ELC values from 2011 to 2019 ranged from 95 to 107.3 with standard deviations ranging from 15.2 to 19.7 - in alignment with the expected mean of 100 and standard deviation of 15. Controlling for differences in age and maternal education via an ANCOVA (**Table 2**), we found inconsistent significant differences in mean ELC scores between 2011-2019 and the second half of 2020, but consistent and significant (p<0.05) reductions between 2011-2019 and 2021. Similar results were found for VDQ and NVDQ. Most differences between 2011-2019 and 2021 remained significant even after strict Bonferonni correction for the 30 tests (corrected p-value < 0.002). Effect sizes (Cohen’s f^2^) for significant differences are listed in the table and range from small (0.05) to large (0.4). The largest effect sizes were noted in the comparisons to the 2021 cognitive values.

**Table 2.** Results of the yearly pairwise comparisons (ANCOVA) in overall, verbal, and non-verbal cognitive scores in all children 0 to 3 years of age. We note near consistent significant reductions in ELC, VDQ, and NVDQ measures in 2021 compared to the past decade (even correcting for multiple comparisons p < 0,002), with less consistent differences in 2020. Effect sizes for significant differences ranged from low (0.05) to high (0.3)

| ReferenceYear | *n* | 2020 ELC | f2 | 2020 VDQ | f2 | 2020 NVDQ | f2 | 2021 ELC | f2 | 2021 VDQ | f2 | 2021 NVDQ | f2 |
| --- | --- | --- | --- | --- | --- | --- | --- | --- | --- | --- | --- | --- | --- |
|  |  | *n=90* |  | *n=90* |  | *n=90* |  | *n=112* |  | *n=112* |  | *n=112* |  |
| 2011 | 110 | 0.002 | 0.057 | <0.001 | 0.051 | 0.012 | 0.036 | <0.001 | 0.164 | <0.001 | 0.123 | <0.001 | 0.138 |
| 2012 | 178 | 0.366 | 0.003 | 0.255 | 0.005 | 0.573 | 0.001 | <0.001 | 0.049 | <0.001 | 0.047 | <0.001 | 0.042 |
| 2013 | 147 | <0.001 | 0.148 | <0.001 | 0.137 | <0.001 | 0.051 | <0.001 | 0.316 | <0.001 | 0.240 | <0.001 | 0.202 |
| 2014 | 82 | <0.001 | 0.276 | <0.001 | 0.192 | <0.001 | 0.151 | <0.001 | 0.406 | <0.001 | 0.273 | <0.001 | 0.360 |
| 2015 | 76 | <0.001 | 0.185 | <0.001 | 0.128 | <0.001 | 0.087 | <0.001 | 0.343 | <0.001 | 0.225 | <0.001 | 0.248 |
| 2016 | 66 | <0.001 | 0.142 | 0.054 | 0.026 | <0.001 | 0.118 | <0.001 | 0.287 | <0.001 | 0.085 | <0.001 | 0.300 |
| 2017 | 62 | <0.001 | 0.053 | 0.025 | 0.038 | 0.154 | 0.015 | <0.001 | 0.169 | <0.001 | 0.111 | <0.001 | 0.108 |
| 2018 | 141 | 0.016 | 0.031 | 0.015 | 0.031 | 0.164 | 0.010 | <0.001 | 0.116 | <0.001 | 0.097 | <0.001 | 0.076 |
| 2019 | 144 | <0.001 | 0.077 | <0.001 | 0.056 | 0.010 | 0.032 | <0.001 | 0.162 | <0.001 | 0.103 | <0.001 | 0.113 |
| 2020a | 39 | <0.001 | 0.065 | <0.001 | 0.088 | <0.001 | 0.762 | <0.001 | 0.110 | <0.001 | 0.092 | <0.001 | 0.085 |

Focussing on infants born before and during the pandemic (**Fig. 1b**), we found similar reductions in performance in 2021 (mean ELC, VDQ, and NVDQ scores of 83.1+/-14.1, 75.5+/-24.1, and 91.1+/-24.1 compared to 2011-2019 (**Table 3**). However, not all comparisons remained significant after correction for the multiple comparisons. Similar effect sizes were noted as found in the larger cohort, ranging from 0.03 to 0.58.

**Table 3.**
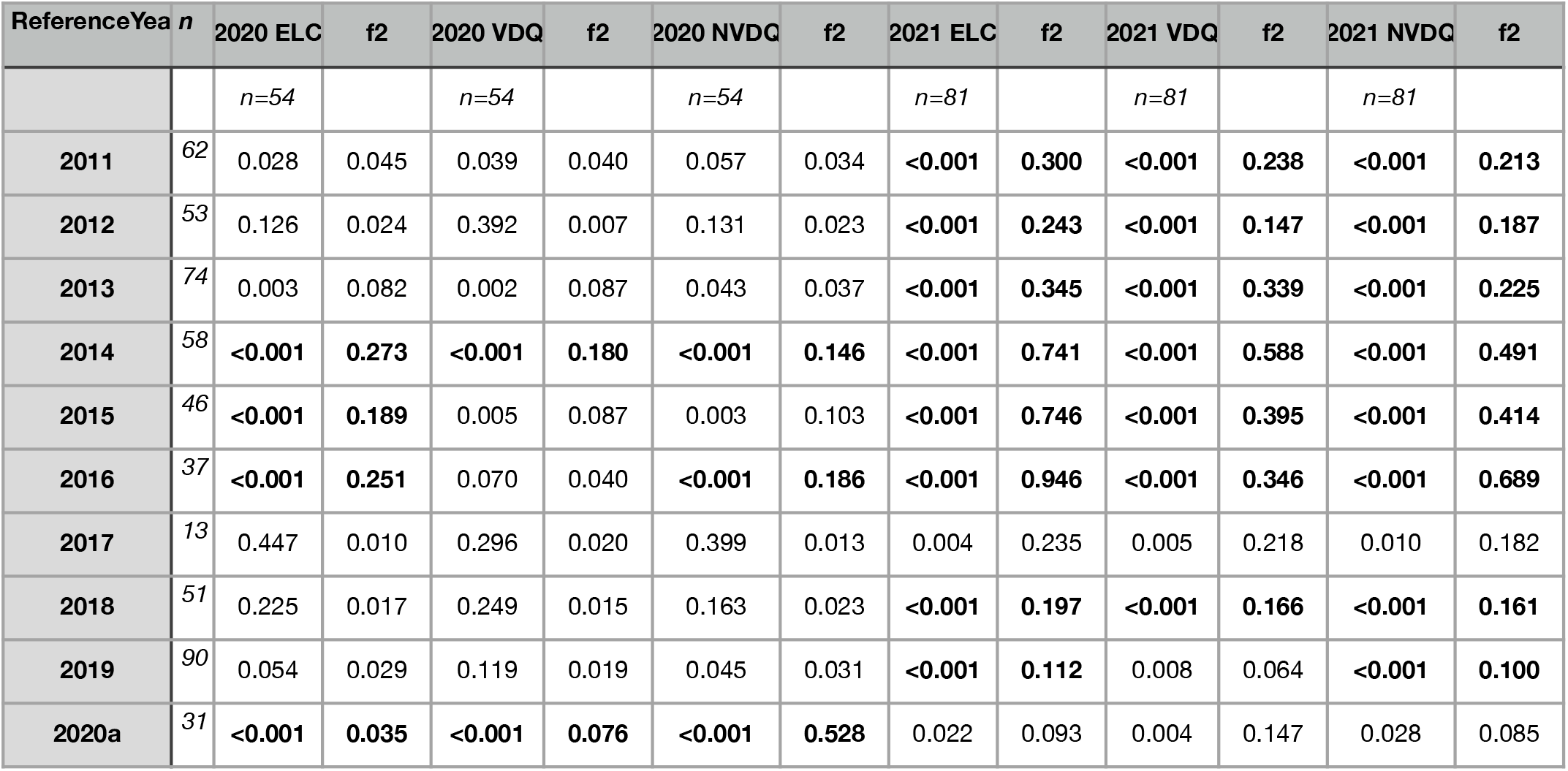
Results of the yearly pairwise comparisons (ANCOVA) in overall, verbal, and non-verbal cognitive scores in infants 0 to 16-months of age. We note near consistent significant reductions in ELC, VDQ, and NVDQ measures in 2020 and 2021 compared to the past decade. Effect sizes for significant differences ranged from low (0.05) to high (0.3)

| ReferenceYear | *n* | 2020 ELC | f2 | 2020 VDQ | f2 | 2020 NVDQ | f2 | 2021 ELC | f2 | 2021 VDQ | f2 | 2021 NVDQ | f2 |
| --- | --- | --- | --- | --- | --- | --- | --- | --- | --- | --- | --- | --- | --- |
|  |  | *n*=54 |  | *n*=54 |  | *n*=54 |  | *n*=81 |  | *n*=81 |  | *n*=81 |  |
| **2011** | <sup>62</sup> | 0.028 | 0.045 | 0.039 | 0.040 | 0.057 | 0.034 | **<0.001** | **0.300** | **<0.001** | **0.238** | **<0.001** | **0.213** |
| **2012** | <sup>53</sup> | 0.126 | 0.024 | 0.392 | 0.007 | 0.131 | 0.023 | **<0.001** | **0.243** | **<0.001** | **0.147** | **<0.001** | **0.187** |
| **2013** | <sup>74</sup> | 0.003 | 0.082 | 0.002 | 0.087 | 0.043 | 0.037 | **<0.001** | **0.345** | **<0.001** | **0.339** | **<0.001** | **0.225** |
| **2014** | <sup>58</sup> | **<0.001** | **0.273** | **<0.001** | **0.180** | **<0.001** | **0.146** | **<0.001** | **0.741** | **<0.001** | **0.588** | **<0.001** | **0.491** |
| **2015** | <sup>46</sup> | **<0.001** | **0.189** | 0.005 | 0.087 | 0.003 | 0.103 | **<0.001** | **0.746** | **<0.001** | **0.395** | **<0.001** | **0.414** |
| **2016** | <sup>37</sup> | **<0.001** | **0.251** | 0.070 | 0.040 | **<0.001** | **0.186** | **<0.001** | **0.946** | **<0.001** | **0.346** | **<0.001** | **0.689** |
| **2017** | <sup>13</sup> | 0.447 | 0.010 | 0.296 | 0.020 | 0.399 | 0.013 | 0.004 | 0.235 | 0.005 | 0.218 | 0.010 | 0.182 |
| **2018** | <sup>51</sup> | 0.225 | 0.017 | 0.249 | 0.015 | 0.163 | 0.023 | **<0.001** | **0.197** | **<0.001** | **0.166** | **<0.001** | **0.161** |
| **2019** | <sup>90</sup> | 0.054 | 0.029 | 0.119 | 0.019 | 0.045 | 0.031 | **<0.001** | **0.112** | 0.008 | 0.064 | **<0.001** | **0.100** |
| **2020a** | <sup>31</sup> | **<0.001** | **0.035** | **<0.001** | **0.076** | **<0.001** | **0.528** | 0.022 | 0.093 | 0.004 | 0.147 | 0.028 | 0.085 |

### Modeling Cognitive Measures over the Past Decade

Results from our series of mixed-model analyses are summarized in **Table 4 (and Supplemental Tables 1 and 2)** and reinforce the ANCOVA findings. For each composite score, the most parsimonious model included child age, SES, child biological sex, birth weight, gestation period, and the PANDEMIC tern, as well as interaction terms of PANDEMIC x biological sex and PANDEMIC x SES (e.g., Eqn. [5]).

**Table 4.**
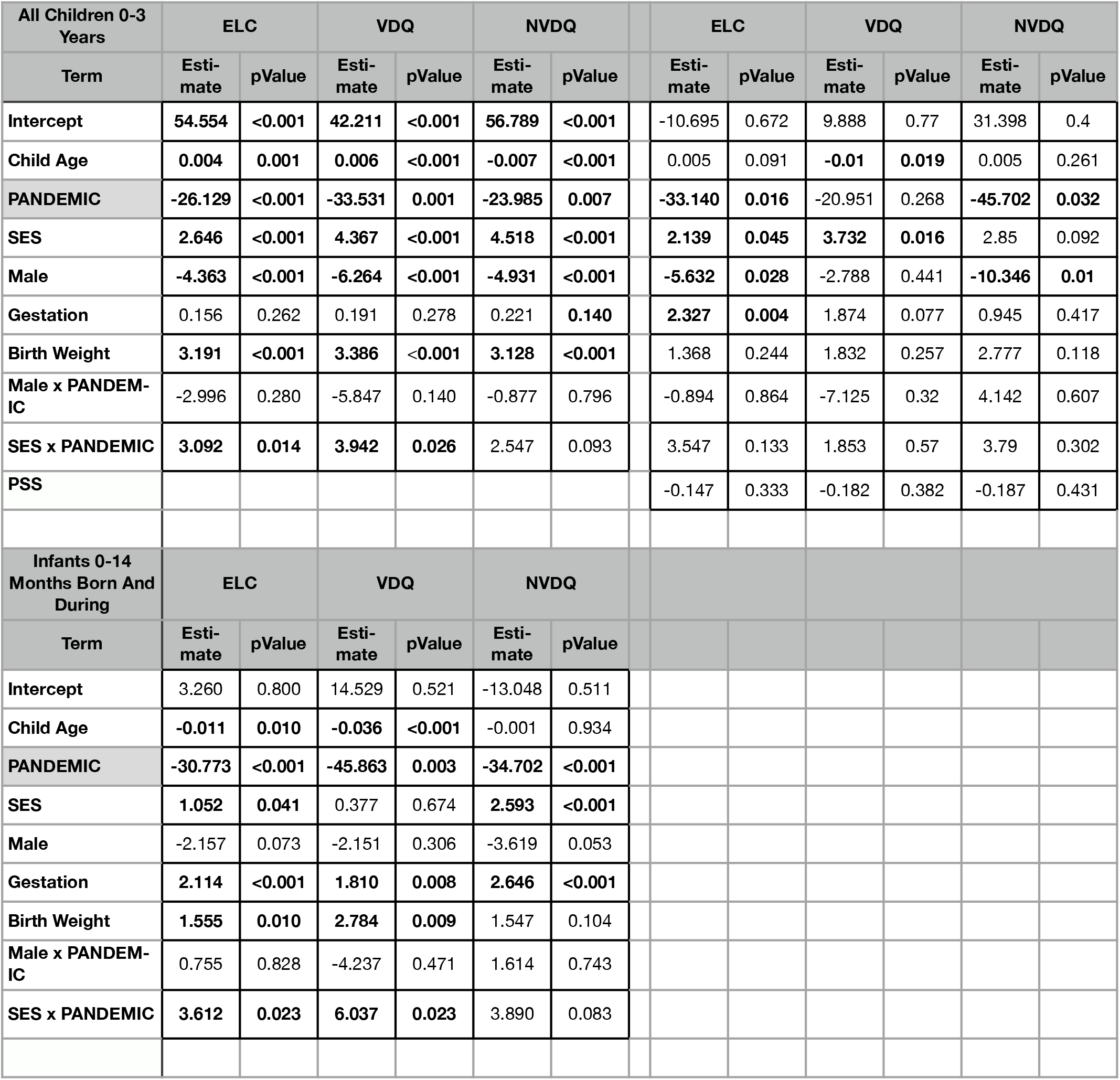
Summary outcomes from our general linear mixed-effects models. In all cases the model including child age, SES, child biological sex, gestation period, birth weight, and the PANDEMIC, PANDEMIC x child biological sex interaction, and PANDEMIC x SES interaction was found to be the most parsimonious. Across all measures in children 0-3 years of age we note that the PANDEMIC term was a significant negative factors, with SES alone and SES x PANDEMIC being significant or trending towards significance. For infants, the PANDEMIC term was a significant negative factor, with increased gestation period and SES x PANDEMIC being a significant positive or protective factor. We also present results of analysis investigating the impact of maternal stress (PSS), which we found to be a non-significant predictor.

We found mean ELC, VDQ, and VDQ scores were significantly reduced by >20 points during the pandemic (or almost two full standard deviations, p < 0.01), with higher maternal education, larger birth weight, and longer gestation duration being protective. Effect size for PANDEMIC term was medium to low, with a f^2^ of 0.73 for ELC, 0.06 for VDQ, and 0.04 for NVDQ.

Performing this analysis in just the infants, we found the same results. Infants born since the beginning of the pandemic exhibit significantly lower cognitive functioning (ELC, VDQ, and NVDQ) compared to infants born over the preceding decade. Gestation, PANDEMIC, and PANDEMIC x SES were also found to be significant predictors across all three main cognitive domains, with increased SES (maternal education) being protective. Again, we found low-to-medium effect size for the PANDEMIC term, ranging from 0.089 for ELC, 0.048 for VDQ, and 0.52 for NVDQ.

### Investigating the Impact of Maternal Stress

One of the hypothesized mechanisms by which the pandemic environment may influence neurodevelopment is through increased maternal stress, both during and following pregnancy. In testing this by either adding the recorded PSS scores to our parsimonious model or including it in place of the PANDEMIC term.

We found that model [6], which included PSS as an additional variable to the general model identified in the above analysis, provided a more parsimonious fit. However, PSS was not a significant predictor in either model (**Table 4** and **Supplemental Table 3**).

## DISCUSSION

Children are inherently shaped by the environment in which they learn, grow, and play. Over the past 20 months, that environment has been fundamentally altered for many children. While the short and long-term neurodevelopmental impacts of the COVID-19 pandemic are not yet well understood in infants and young children (49), emerging reports are suggestive of reduced motor and communication skills (38). Results from this study add to this literature and are in agreement with these past studies. We find that verbal, non-verbal, and overall cognitive scores in children under 3 years of age are significantly lower than over the past decade with this difference amplified in infants <16-months of age and born since the beginning of the pandemic. As the pregnant individuals and children included in this study reported no symptoms of SARS-CoV-2 infection or had positive antibody or RT-PCR test results, this suggests that observed effects are related to environmental factors rather than due to direct effects of infection. However, we did not perform independent antibody testing to confirm past infection status.

Longitudinal modeling of children 0-3 years of age revealed positive associations between child cognitive performance and maternal education, gestation duration, and birth weight; and negative associations with male biological sex. In addition, we found a positive association between cognitive scores and the interaction of maternal education x PANDEMIC. Socioeconomic characteristics such as material education have well-known associations with child health and cognitive development and may be related to differences in the ability to provide adequate nutrition, supportive parenting, and stimulating and engaging environments. That increased maternal education was found to buffer the negative effects of the pandemic may be related to these families being able to provide continued stability (lower food and/or housing insecurity) and interactive stimulation (e.g., reading and play). In contrast, many lower-income families have dealt with job loss and financial insecurity that has necessitated taking on additional work and reducing the available family time and/or ability to provide quality daycare. Additional investigation of changes in food and housing insecurity, shifts in daycare, and employment status throughout the pandemic is needed to understand these individual factors but was unavailable here.

Other significant factors identified throughout our analysis were gestation length and infant birth weight. While this aligns with past studies of infant birth outcomes and subsequent development, most have focused on premature and low birth weight infants - infants excluded from our analysis. However, even amongst healthy birthweight infants, there is evidence that increased birth weight and later math and language processing (50). Similarly, even within children born full-term, later infant and children neurocognitive outcomes have been shown to improve for each week of gestation past 37 weeks, with a peak at 40-41 weeks (51).

In contrast, we found that males have overall lower neurodevelopment. The reason for this finding in our cohort is unclear, however, past analysis of male and female cognitive development using the MSEL has shown that in the context of children of mothers with depression, females performed better than males and improved more quickly between 6 and 18-months of age (52), overlapping with the age range investigated here.

One hypothesized mechanism by which the environment may affect neurocognitive development is via maternal pre-and postnatal stress. However, in contrast to other studies of mothers during the pandemic, such as the MOM-COPE study and a large online survey of pregnant individuals at the beginning of the pandemic (53, 36), we did not observe increased maternal stress in our study population, and it was not a significant predictive factor in our analysis. Two possible explanations for this are 1. Insensitivity of the PSS tool used to pandemic-specific stress; and 2. A potential selection bias in the families included in our study. Addressing these explanations, in turn, the PSS is a standardized ten-item questionnaire that asks about general life stressors and how stressful individuals find their lives. It, however, does not include specific questions related to health or wellbeing. In contrast, the MOM-COPE study utilized retrospective data collection using an ad-hoc developed questionnaire focused on worry and anxiety of COVID-19 infection, pregnancy risk, and their own and their infant’s health (36).

The online pregnancy survey study by Lebel and colleagues (53) also used a specially developed questionnaire to gauge maternal concern of the pandemic and its impact on their own and their infant’s health. Thus, it is possible that stress felt by parents and families specific to COVID-19 infection or complications during pregnancy was not identified on the PSS given its generic and broad nature that may have been revealed with more sensitive tools.

A second explanation as to why the stress was not found to be a significant factor is the potential selection bias of the families included in this study and, in particular, those who were assessed during the pandemic. Unlike many other academic research laboratories across the US, our facility has continued to operate throughout the pandemic with in-person study visits, with appropriate pre-screening and masking policies. As our center is located within a clinical setting, parents less concerned about the pandemic may have been more likely to participate than those with greater concerns. Thus, our observation that maternal stress (PSS) was not significantly increased may simply reflect the reality that we only tested less stressed and anxious families.

In the absence of stress-related changes, additional factors that may explain our findings are the use of face masks during testing. Although all study visits were performed in-person, the inability of infants to see full facial expressions may have eliminated non-verbal cues, muffled instructions, or otherwise impair the understanding of test questions and instructions. Without direct comparison of performance in the same children with and without face masks, it is difficult to rule in or out the potential influence of masks. However, some insight can be gleaned by examining the raw domain scores (**Supplemental Figure 3**). Over the first year of a child’s life, the MSEL focuses strongly on non-verbal domains such as 1. Gross motor control skills: e.g., supporting their head, pushing up on their forearms, rolling over, and pulling themselves up to standing; 2. Fine motor skills: e.g., holding rings and grasping and reaching for objects; and 3. Visual skills: e.g., fixation and face tracking. Tested verbal skills are also relatively basic, including orienting to sounds, responding to a voice, swallowing, making coos, chuckles, or laughing sounds, and babbling. For much of the assessment, the examiner is on the floor playing with and manipulating the child, and the assessment is not reliant on only verbal cues or instructions. In examining raw domain scores, we note that overall infants born since the pandemic are most affected in their basic motor and visual receptive skills rather than their expressive and receptive language abilities. Given the nature of these affected skills, it seems unlikely that mask-wearing by the experimenter could be the cause of the reduced scores in late 2020 and 2021. Of note, recent work using parent-reported development measures reported similar findings of reduced motor development in 6-month old infants born during the pandemic, further suggesting masks do not underly our observed findings.

Secondary to masks, however, may be the unfamiliarity of infants and toddlers to individuals outside of their immediate family. Infants may have been distracted in the unfamiliar laboratory setting or suffered “performance anxiety” in front of the unknown research staff and testers testing strangers even with their parents nearby. Our research staff and neuropsychologists are highly trained and well versed in interacting with children across this age group and working with the child and their parents to obtain the most representative assessment.

Our findings raise important questions. First, do they generalize across the US or are they specific to the RI population and the cohort investigated? Despite the relatively restricted sample size, particularly within the infant cohort, we note the high effect sizes in our findings suggest our findings should be reproducible. Indeed, our results align with other emerging reports from other cohorts that used different assessment tools. It is unclear how different state-level responses to the pandemic will affect child outcomes, for example, Arkansas, Iowa, Nebraska, North Dakota, South Dakota, Utah, and Wyoming that did not issue stay-at-home orders. Second, are observed differences in infant neurocognitive performance since the pandemic temporary, and will they normalize as children grow and return to prepandemic levels of play, interaction, schooling, etc. Unfortunately, the appearance of new virus variants and continued high infection and hospitalization rates have prolonged the much-anticipated return to “normal”. Our data, currently, is not sufficient to determine if observed reductions are transient or longer-lasting. However, the longitudinal nature of our study will allow us to address this in time as individuals return. It is clear, however, that young infants and children are being affected now. Programs such as unemployment insurance, Supplemental Nutrition Assistance Program (SNAP), the Special Supplemental Nutrition Program for Women, Infants, and Children (WIC), and housing assistance, may help minimize the impact of the pandemic on the most sensitive of children. In addition, further research directly exploring aspects of parent-child attachment, interaction, nutrition, food security, and environmental stimulation is needed to understand the primary causative factors underlying the results presented here.

## CONCLUSION

The COVID-19 pandemic has fundamentally altered the child health landscape, with families living in strikingly different economic, psychosocial, and educational environments than what was present just 18 months ago. Against this environmental backdrop, unanswered questions remain regarding the impact of the work-from-home, shelter-in-place, and other public health policies that have limited social interaction and typical childhood experiences on early child neurodevelopment. In this work, we provide early evidence suggestive of significant reductions in attained cognitive function and performance in children born over the past 18 months during the pandemic. While maternal education appears to mitigate against the negative consequences of the pandemic, the primary factors underlying this observation remain unclear. Understanding these factors is critical to helping ensure affected children rebound as the pandemic winds down and they re-enter daycares and schools; as well as implementing additional public health and educational policies that address the most affected of children, particularly those in lower-income families.

## Data Availability

All data used in this study are available in de-identified form upon request.

## List of Abbreviations

ELC: Early Learning Composite
MSEL: Mullen Scales of Early Learning
NVDQ: Non-Verbal Development Quotient
VDQ: Verbal Development Quotient

## DATA SHARING

All data acquired and presented here is available upon request to the authors.

## CONTRIBUTOR ROLES

All listed authors were involved in the study design, data acquisition, and analysis, drafting and revising this manuscript, and providing final and accountable approval for its contents. SCD and DV verify the underlying data.

## Supplemental Table Legends

**Supplemental Table 1.**
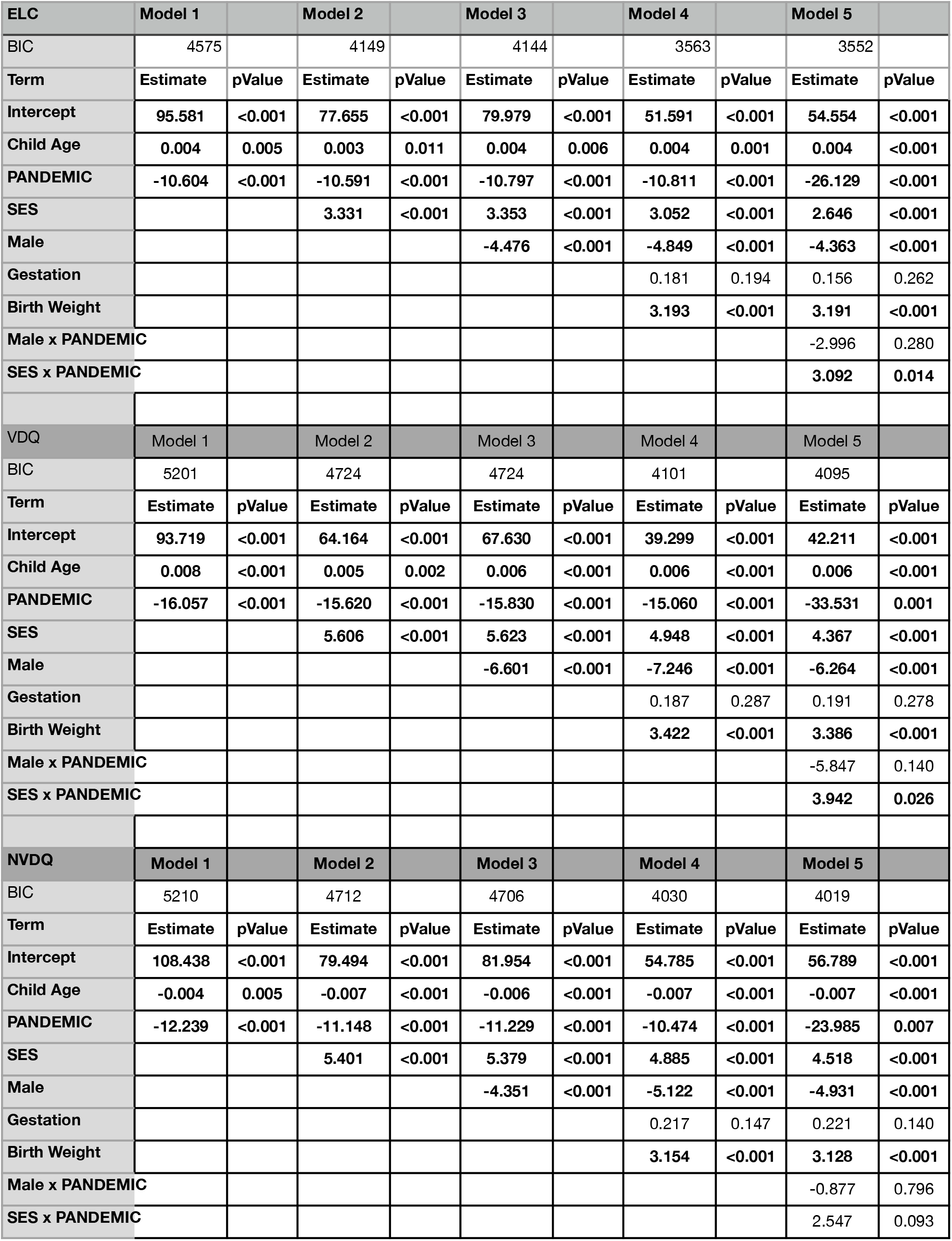
Complete results from our step-wise general linear mixed-effect model analysis for all children, 0 to 3 years of age.

**Supplemental Table 2.**
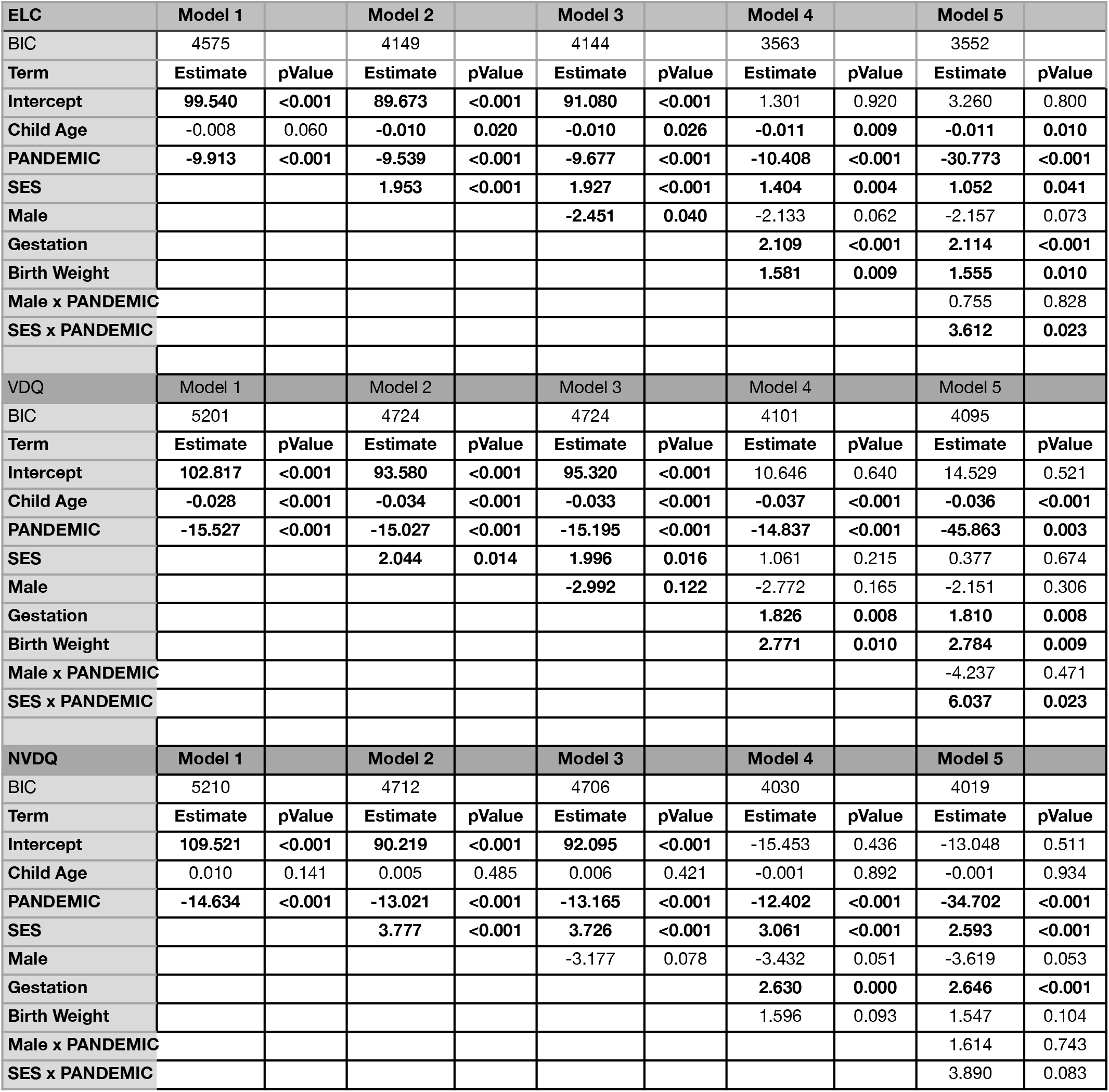
Complete results from our step-wise general linear mixed-effect model analysis for all children, 0 to 16 months of age.

**Supplemental Table 3.** Complete results from our general linear mixed-effect model analysis examining the impact of maternal stress.

| ELC | Model 1 |  | Model 2 |  |
| --- | --- | --- | --- | --- |
| Term | Estimate | pValue | Estimate | pValue |
| Intercept | -2.803798 | 0.923559 | -10.695 | 0.672 |
| Child Age | 0.002642 | 0.412242 | 0.005 | 0.091 |
| **PANDEMIC** |  |  | **-33.14** | **0.016** |
| SES | 4.201352 | **0.036024** | **2.139** | **0.045** |
| Male | -8.879188 | 0.084166 | **-5.632** | **0.028** |
| Gestation | 1.876727 | **0.026806** | **2.327** | **0.004** |
| Birth Weight | 1.162756 | 0.358609 | 1.368 | 0.244 |
| Male x PANDEMIC |  |  | -0.894 | 0.864 |
| SES x PANDEMIC |  |  | 3.547 | 0.133 |
| PSS | -0.058189 | 0.933868 | -0.147 | 0.333 |
| Male x PSS | 0.133976 | 0.68375 |  |  |
| SES x PSS | -0.042414 | 0.732502 |  |  |
| **VDQ** | **Model 1** |  | **Model 2** |  |
| Term | Estimate | pValue | Estimate | pValue |
| Intercept | 16.634546 | 0.701326 | 9.888 | 0.77 |
| Child Age | 0.003645 | 0.462384 | -0.01 | 0.019 |
| **PANDEMIC** |  |  | -20.951 | 0.268 |
| SES | 8.949226 | **0.003374** | 3.732 | 0.016 |
| Male | -13.790175 | 0.081917 | -2.788 | 0.441 |
| Gestation | 0.467872 | 0.702321 | 1.874 | 0.077 |
| Birth Weight | 2.409064 | 0.202964 | 1.832 | 0.257 |
| Male x PANDEMIC |  |  | -7.125 | 0.32 |
| SES x PANDEMIC |  |  | 1.853 | 0.57 |
| PSS | 1.143643 | 0.298257 | -0.182 | 0.382 |
| Male x PSS | 0.206981 | 0.683043 |  |  |
| SES x PSS | -0.273539 | 0.155058 |  |  |
| **NVDQ** | **Model 1** |  | **Model 2** |  |
| Term | Estimate | pValue | Estimate | pValue |
| Intercept | 16.129832 | 0.672381 | 31.398 | 0.4 |
| Child Age | -0.010764 | **0.013355** | 0.005 | 0.261 |
| **PANDEMIC** |  |  | **-45.702** | **0.032** |
| SES | 4.870884 | 0.065612 | 2.85 | 0.092 |
| Male | -10.533211 | 0.126408 | **-10.346** | **0.01** |
| Gestation | 1.650778 | 0.128961 | 0.945 | 0.417 |
| Birth Weight | -0.475995 | 0.613823 | 2.777 | 0.118 |
| Male x PANDEMIC |  |  | 4.142 | 0.607 |
| SES x PANDEMIC |  |  | 3.79 | 0.302 |
| PSS | 1.468026 | 0.382358 | -0.187 | 0.431 |
| Male x PSS | 0.363321 | 0.406477 |  |  |
| SES x PSS | 0.010763 | 0.948116 |  |  |

## Supplemental Figure Legends

**Supplemental Figure 1.**
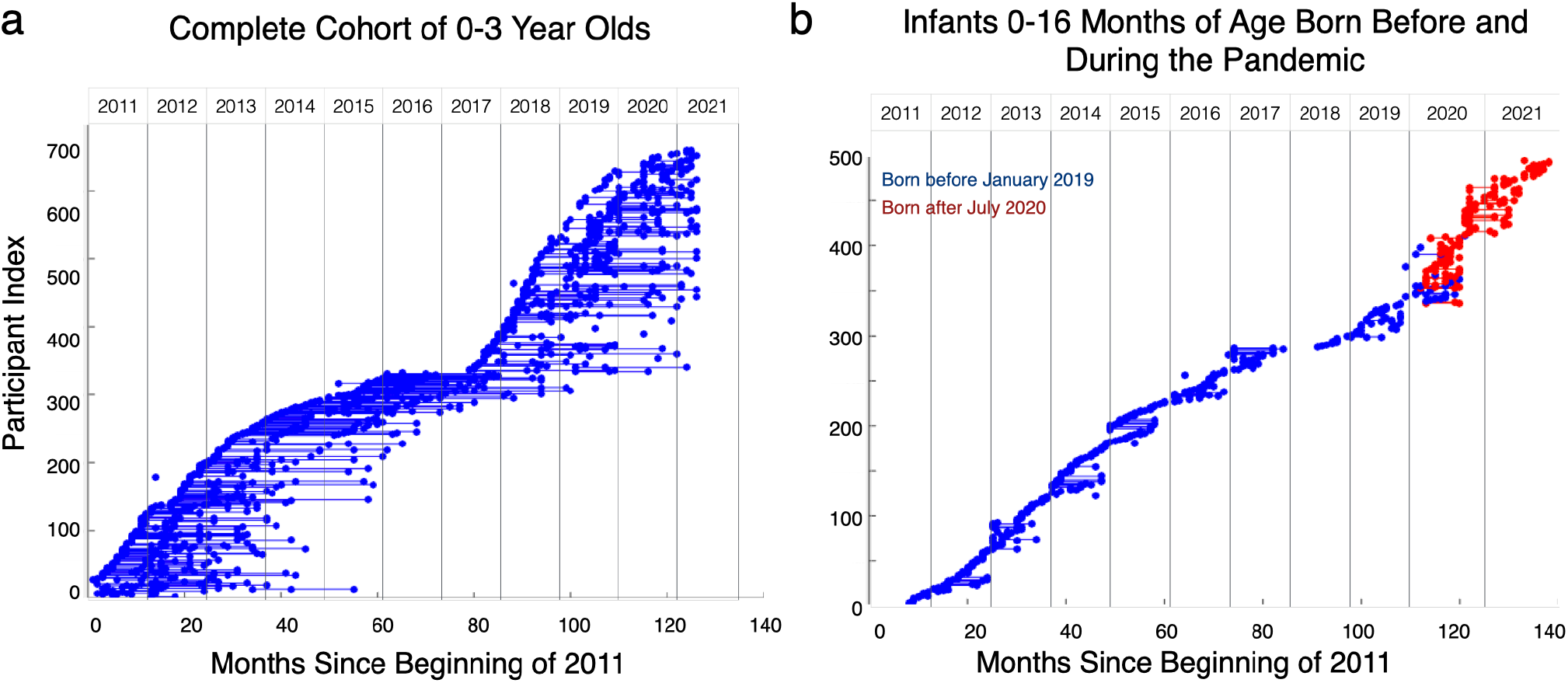
Visual overview of all child study visits used in each set of analysis. (a) All children from 0 to 3 years of age; (b) Infants born prior to (blue) or during (red) the pandemic.

**Supplemental Figure 2.**
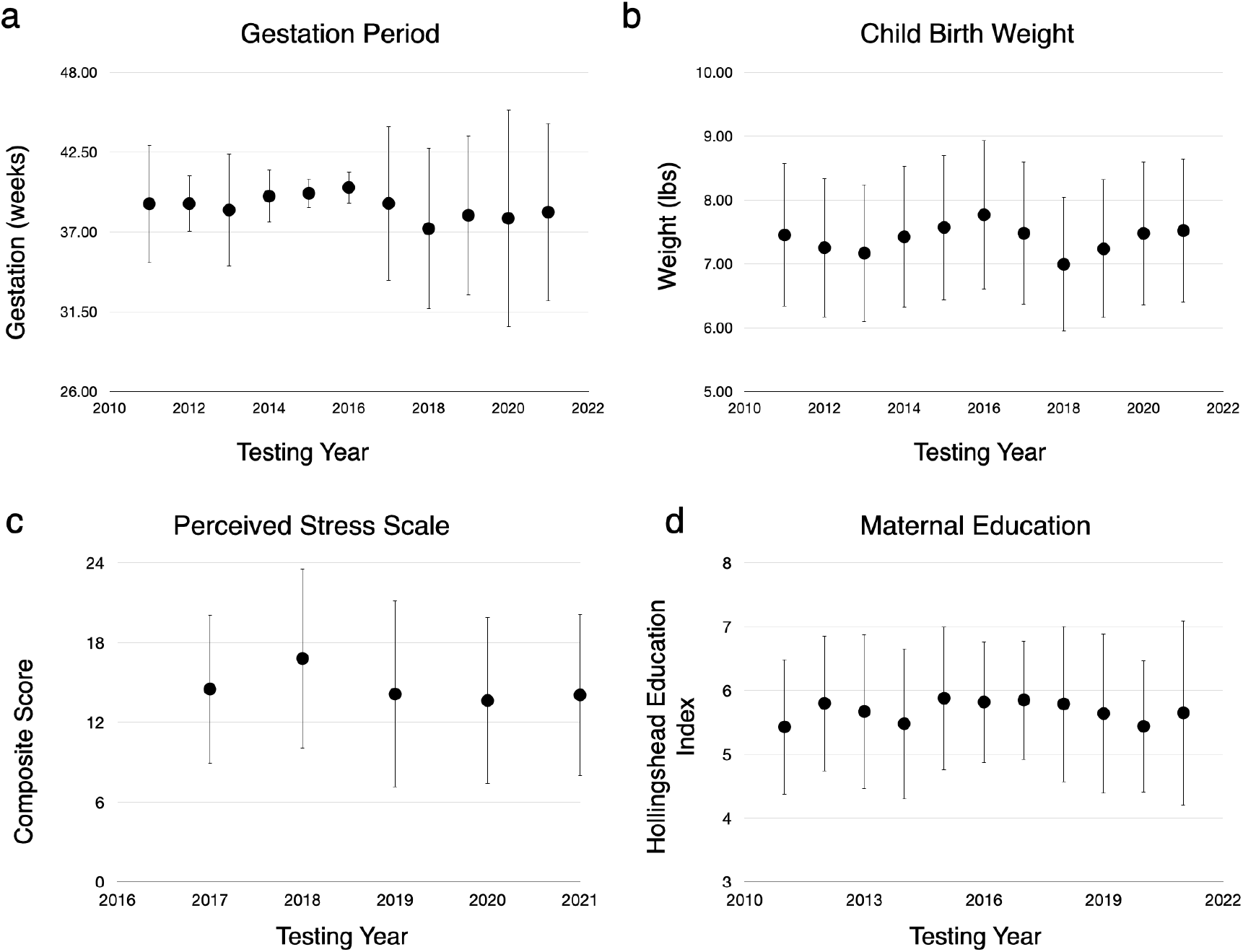
Plots of the mean gestation period, birth weight, maternal education, and maternal perceived stress for our child sample that were measured each year.

**Supplemental Figure 3.**
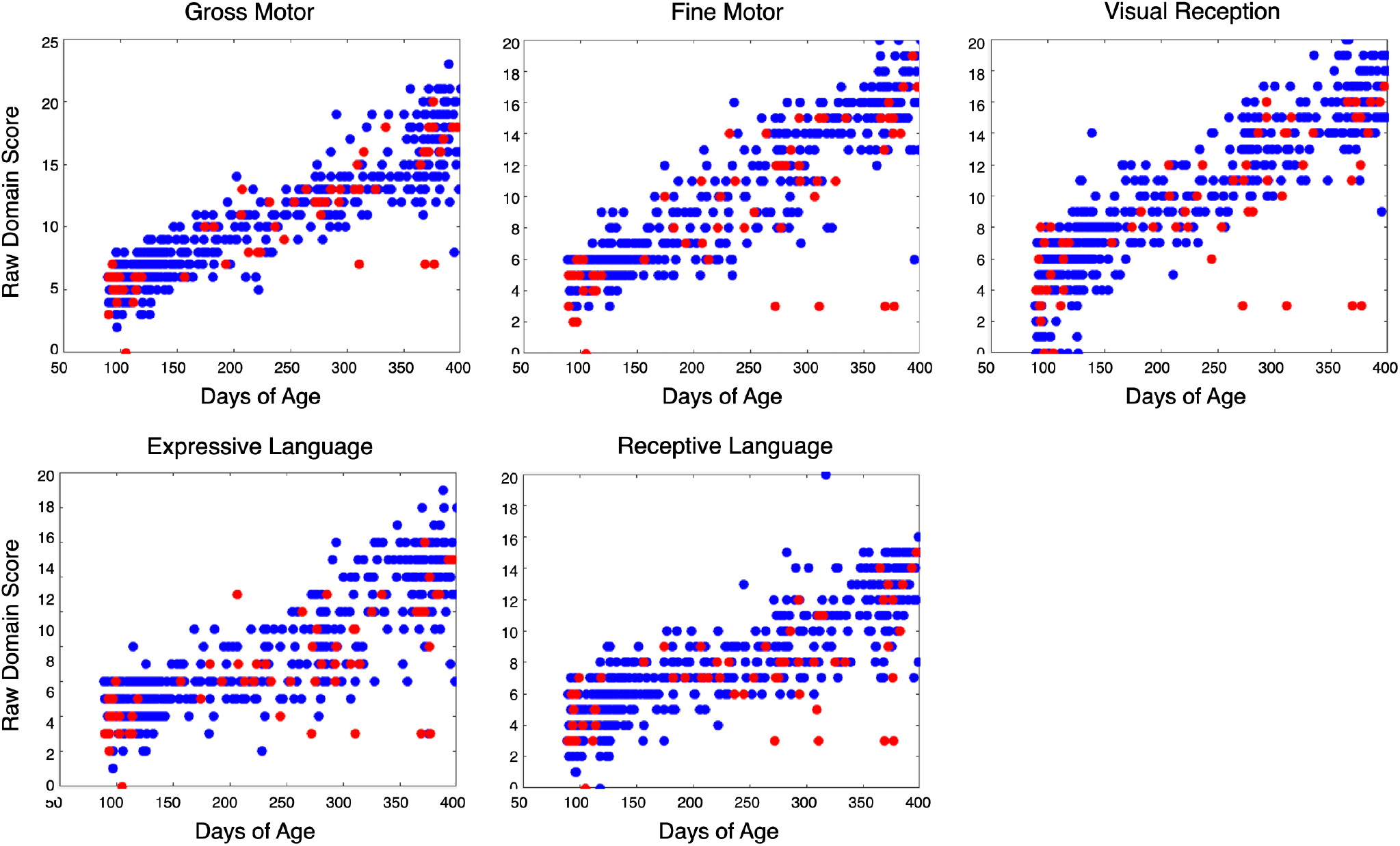
Raw MSEL domain scores in 0 to 16-month-old infants born before the pandemic (blue) and during the pandemic (red).

